# The neurostructural consequences of glaucoma and their overlap with disorders exhibiting emotional dysregulations: a voxel-based meta-analysis and tripartite system model

**DOI:** 10.1101/2023.10.11.23296863

**Authors:** Benjamin Klugah-Brown, Mercy C. Bore, Xiqin Liu, Xianyang Gan, Bharat B. Biswal, Keith M. Kendrick, Dorita Chang, Bo Zhou, Benjamin Becker

## Abstract

Glaucoma is a neurodegenerative disorder characterized by progressive optic nerve damage and the most common cause for irreversible blindness. Emotional dysregulations are common in glaucoma patients, with high rates of generalized anxiety and depression. Glaucoma leads to pathological changes in the visual pathways of the brain, yet accumulating evidence suggests that the brain changes may reach beyond the visual system. To robustly determine brain morphological alterations in glaucoma and determine a potential overlap with changes observed in anxiety and depression we performed a pre-registered comparative meta-analysis of case control studies examining brain structural integrity in patients with glaucoma, and further determined whether the identified regions are nodes of distinct overarching large-scale networks and overlap with meta-analytic brain structural maps of generalized anxiety disorder (GAD) and major depressive disorder (MDD). Glaucoma patients exhibited robust reductions in gray matter volume in regions contributing to visual processing (lingual gyrus, thalamus), but volumetric reductions extended beyond systems involved in visual processing and also affected the left putamen and insula. Behavioral and functional network level decoding demonstrated distinct large-scale networks and involvement in visual, motivational or affective domains, respectively. Reductions in the insular region involved in pain and affective processes overlapped with alterations previously observed in GAD. Our findings may suggest a tripartite brain model of glaucoma, with changes in visual processing regions such as the lingual gyrus as well as additional alterations in putamen and insular regions meta-analytically associated with motivational or emotional functions, respectively. Together the findings indicate broad neuroanatomical alterations in glaucoma that extend beyond the visual system and may gate further pathological developments.

## Introduction

Glaucoma is a neurodegenerative disorder characterized by a progressive damage of the optic nerve, leading to a gradual loss of vision and eventually irreversible blindness (Tham et al., 2014). With an estimated 79.6 million individuals affected by glaucoma in 2020 the disorder has become the leading cause of irreversible blindness worldwide (Quigley & Broman, 2006). Accumulating evidence suggests that the disorder is accompanied by neuropathological alterations that affect the integrity of the brain beyond the optic nerve and the primary visual system (Nuzzi et al., 2018), however, the extent of the brain-based alterations has not been robustly determined. Recent surveys moreover reported a high prevalence of stress– and emotion-related mental disorders in patients with glaucoma (Nuzzi et al., 2018). Against this background, it is imperative to robustly determine the neuroanatomical changes that may accompany or even underlie the visual loss but also to determine alterations potentially related to the emotional dysfunctions.

The most common type of glaucoma is open-angle glaucoma (Hernandez, 1992), characterized by increased intraocular pressure that progressively damages the optic nerve responsible for transmitting visual information to the brain (Weinreb et al., 2014). Although elevated intraocular pressure is a well-known major risk factor for glaucoma, the condition often leads to progressive vision loss despite efficacious interventions that reduce intraocular pressure (Boland & Quigley, 2007). These observations indicate that factors other than intraocular pressure may also play a role in the pathological progression of the disease (Gupta & Yücel, 2007). Mounting evidence indicates that neurodegeneration represents a significant pathogenic and progression mechanism in glaucoma (Gupta et al., 2006), and various neuroprotective agents are currently being investigated for their potential therapeutic potential in glaucoma.

In recent years, brain imaging techniques in particular brain structural magnetic resonance imaging (MRI) have been utilized to better determine the neurostructural alterations in glaucoma. The majority of the studies to date have employed voxel-based morphometry (VBM) techniques which allow an automated detection of regional alterations in brain structure while allowing avoidance of a bias toward any particular brain region (Astrakas & Argyropoulou, 2010). In line with the primary visual symptoms of the disease, neurostructural alterations have been most commonly observed in the primary and secondary visual processing systems of the occipital cortex ((Boucard et al., 2009; Frezzotti et al., 2014) see overview in (Nuzzi et al., 2018)), yet an increasing number of studies reported neurostructural alterations that reach beyond the core visual processing systems and encompass limbic and striatal regions (e.g (Wang et al., 2020)). Variations in the structural integrity of these limbic and striatal regions have repeatedly been associated with emotional and motivational dysregulations, including elevated anxiety and depression (Gray et al., 2020; Klugah-Brown et al., 2021, 2022; C. Liu et al., 2019; X. Liu et al., 2022; Serra-Blasco et al., 2021) and may render individuals at a greater risk for the development of emotional dysregulations (Becker et al., 2015; Talati et al., 2022).

Although the increasing number of brain structural studies in glaucoma provide accumulating evidence for a neuroanatomical basis of the disease, findings remained inconsistent and of limited generalizability due to the limitations inherent to the original case-control studies including a small sample size and heterogeneity in the patients as well as heterogeneity in the data processing methods for VBM (X. Zhou et al., 2022). Coordinate-based meta-analysis (CBMA) techniques for neuroimaging data have been developed allowing to integrate findings from multiple studies and to establish the development of robust brain markers for mental and neurological disorders (Eickhoff et al., 2016; Laird et al., 2005; Tench et al., 2017; Turkeltaub et al., 2012; Wager et al., 2007). CBMA allows a quantitative weighing of the evidence from previous brain imaging studies and to determine brain regions that exhibit convergent alterations by integrating data from previous published brain imaging studies. This approach strongly enhances the statistical power and reproducibility of the findings. By integrating findings from multiple VBM in studies in glaucoma patients, CBMA thus allows a more comprehensive and reliable understanding of the neuroanatomical changes associated with glaucoma. The approach additionally allows determining for the first time common and separable neurostructural alterations with stress-related disorders that are highly common in these patients, in particular depression and generalized anxiety thus allowing an exploration of brain systems that potentially mediate emotional dysregulations in glaucoma patients.

To this aim we implemented a CBMA that followed state of the art protocols, including preregistration of procedures and hypotheses and the Preferred Reporting Items for Systematic Reviews and Meta-Analyses (PRISMA) guidelines (Parums, 2021). The literature search utilized comprehensive biomedical and life science databases, including PubMed and Web of Science. By capitalizing on CBMA we next identified robust brain structural changes in glaucoma and explored potential overlap with structural alterations in mental disorders that show a particular high co-morbidity with glaucoma, i.e., major depression (MDD) and generalized anxiety disorder (GAD) (Rezapour et al., 2018). We employed the Seed-based d Mapping with Permutation of Subject Images (SDM-PSI) (Albajes-Eizagirre, Solanes, Fullana, et al., 2019) technique, a robust and novel CBMA method that allows the implementation of disorder-specific and comparative conjunction meta-analyses. These analyses aimed to identify disorder-specific brain structural alterations in glaucoma and to compare them with established meta-analytic maps for brain structural alterations in anxiety and depression (as determined in our previous CBMA meta-analysis in depression and anxiety, see (X. Liu et al., 2022)). In addition to identifying structural alterations the study employed meta-analytic network level and behavioral decoding techniques to characterize and further segregate the identified alterations on the system level.

## Methods

The present study was pre-registered on the OSF-repository (https://osf.io/ay3r9) and followed the guidelines for conducting coordinate-based neuroimaging meta-analyses (Müller et al., 2018). Procedures, hypotheses, and analyses were pre-registered. Biomedical and life science databases, including PubMed (https://www.ncbi.nlm.nih.gov/pubmed) and Web of Science (https://www.webofscience.com/wos/alldb/basic-search), were utilized for the literature search.

Additionally, we included relevant studies from the reference lists of review articles. The primary search aimed at identifying suitable case-control studies that examined brain structural alterations in glaucoma patients using MRI in combination with VBM. The inclusion criteria were as follows: studies written in English, published between the beginning of the study coverage and June 2022, and reporting whole brain results. The literature was screened based on titles and abstracts obtained from search results using the following search terms: “Magnetic Resonance Imaging” OR “MRI” OR “structural changes” AND “visual” OR “glaucoma” OR (glaucom* AND disease) OR POAG OR NTG OR PACG) AND “gray matter” OR “Voxel-based morphometry” OR “VBM” OR “voxel-wise”. The comparative brain structural maps for depression and anxiety were from our previously published brain structural meta-analysis that synthesized VBM results from case-control studies in depression (MDD) and generalized anxiety disorder (GAD) using comparable search procedures and meta-analytic techniques (details see (X. Liu et al., 2022)).

## Aims and coordinate-based meta-analytic implementation

The present case-control neuroimaging meta-analysis aimed to determine (1) robust brain structural alterations as identified via structural MRI in patients with glaucoma as compared to healthy controls (HC), (2) characterize the identified regions on the behavioral and network level, and (3) determine a potential overlap between the identified neurostructural alterations in glaucoma with brain structural alterations in patients MDD and GAD as determined in our previous meta-analysis (X. Liu et al., 2022).

All analyses were implemented in SDM-PSI version 6.21 (https://www.sdmproject.com/) a novel and highly robust technique for performing neuroimaging meta-analyses (Albajes-Eizagirre, Solanes, Fullana, et al., 2019). In line with the aims of the study we implemented a three-step meta-analytic approach, including (1) a single-disorder meta-analysis that pooled data from published VBM case-control studies in glaucoma patients to determine robust GMV alterations in glaucoma, (2) meta-analytic co-activation, connectivity and behavioral decoding analyses to characterize the network level communication and general behavioral functions of the identified regions, (3) conjunction meta-analyses that compared neurostructural alterations in glaucoma with those reported in our previous study on MDD and GAD (X. Liu et al., 2022) and that accounted for the error in the estimation of *p*-values within each voxel from the separate meta-analytic maps.

## Neuroimaging meta-analysis and thresholding

The SDM-PSI approach followed evaluated protocols: (a) Peak coordinates of between-group differences and their effect sizes in terms of either t/z values were extracted according to the SDM software procedures of conducting a meta-analysis (Albajes-Eizagirre, Solanes, & Radua, 2019). Z-values representing coordinates from whole-brain between-group differences of depressed patients and healthy controls were transformed into t-values using the statistical converter (https://www.sdmproject.com/utilities/?show=Coordinates). (b) For studies reporting peak coordinates, the initial preprocessing step (Anisotropic full width half maximum (FWHM) = 20mm and voxel size = 2mm) estimated the lower and upper bounds (Hedges’ g) of the most probable effect size images (Radua et al., 2014). (c) The mean is analyzed by the maximum-likelihood estimation (MLE) and meta-analysis of non-statistically significant unreported effects (MetaNSUE) algorithm. This not only estimates the most likely effect size and the standard error, but it also creates numerous imputations based on the estimates that are within the bounds (Albajes-Eizagirre, Solanes, & Radua, 2019). (d) Next, imputed study images are recreated. (e) Finally, the permutation test evaluated the combined meta-analysis images for statistical significance. The generated maps were visualized using multi-image analysis GUI (MANGO; http://ric.uthscsa.edu/mango). Meta-analytic thresholding was performed using p<0.0025 uncorrected, k > 10 (in line with previous studies e.g. X. Liu et al., 2022).

## Sensitivity, heterogeneity and publication bias

A series of analyses was employed to test the robustness of the identified regions in glaucoma. Inter-study heterogeneity was examined by evaluating the *I^2^* index for each cluster. The index represents the proportion of the total variation caused by study heterogeneity (Higgins & Thompson, 2002), with *I^2^*> 50% suggesting substantial heterogeneity (Martins & Paduraru, 2022). Robustness of the results was further tested using a Jackknife sensitivity analysis as implemented in Anisotropic Effect Size-Signed Differential Mapping (AES-SDM). Funnel plots and Egger’s tests were further employed to explore publication bias.

## Characterization of networks and on the behavioral level

First, we utilized meta-analytic network analyses to characterize the identified regions on the system level and to determine whether they represent nodes of separable networks. Functional decoding was conducted using the Neurosynth database (Yarkoni et al., 2011) (https://www.neurosynth.org/). The peak coordinates of the identified regions were used to generate region-specific unthresholded resting-state functional connectivity maps, meta-analytic co-activation maps, and their respective conjunction maps. The resulting resting-state functional connectivity maps were thresholded at r > 0.2 and transformed into z-scores. Next, conjunction maps between the thresholded resting-state functional connectivity and meta-analytic co-activation maps were generated for each ROI signature using SPM, and the resultant regions were identified with the Anatomy toolbox (Eickhoff et al., 2005, 2006, 2007). The functional connectivity maps represent brain regions co-activated across the resting-state fMRI time series with the seed regions. Meta-analytic co-activation maps symbolize the co-activation of brain regions across all fMRI studies in the Neurosynth database. The combination of functional connectivity and meta-analytic co-activation maps allows analysis of both task-independent and task-driven functional networks emerging from the seed region. Second, distinct behavioral functions of the regions were identified through meta-analytic topic mapping using the peak coordinates of the identified brain regions obtained from the Neurosynth decoder. The top 10 behavioral terms, along with their p-values, were obtained and displayed.

## Results

### Included studies and sample characteristics

The primary search identified glaucoma (n=8) studies that adhered to our inclusion criteria, while the comparative meta-analyses are based on data from GAD (n=9) studies, and MDD (n=46) studies respectively as identified in (X. Liu et al., 2022). Table 1 provides the general information of the glaucoma studies and Table 2 shows demographic and clinical information of the individual studies. The meta-analysis included patient data for glaucoma (n=207, mean age=52.47, *SD*=10.59), GAD (n=226, mean age=28.90, *SD*=7.02) and MDD (n=2575, mean age=36.49, *SD*=9.11). Further, data from healthy controls was included for glaucoma (n= 174, mean age=44.63, *SD*=7.55), GAD (n=226, mean age=28.62, SD=7.44) and MDD (n=2866, mean age=34.66, *SD*=8.85). There were no significant between-group differences in age (p=0.28, t=2.14) and gender in the glaucoma group (p=0.94, t=2.14). There were no significant between-group differences in age (p=0.71, F=7.71) and gender (p=0.99, F=7.70) across all disorders.

**Table 1.** General information of the 8 included studies of glaucoma.

| Authors | Type of Glaucoma | Unilateral/Bilateral | Comorbidity Y/N? | Significant GM alterations | Ocular characteristics |
| --- | --- | --- | --- | --- | --- |
| Li et al. 2022 | NTG | Bilateral | No | Decreased GM volume | NA |
| Wang et al. 2020 | HTG | Unilateral and Bilateral | No | Decreased GM volume | NA |
| Li et al. 2012 | POAG | Bilateral | No | Decreased GM density | Mean defect+ Early stage = $-2.48 \pm 2.04$(right eye), $-2.51 \pm 1.67$(left eye). Advanced/Late stage POAG= $-13.65 \pm 7.79$(right eye), $-16.75 \pm 12.32$(left eye). |
| Chen et al. 2013 | POAG | Bilateral | No | Increased and decreased GM volume | Visual acuity, LogMAR= right eye, $0.15 \pm 0.13$ and left eye, $0.16 \pm 0.15$. IOP before treatment= right eye, $28.1 \pm 3.9$, left eye, $30.1 \pm 4.3$ |
| Zikou et al. 2012 | POAG | Bilateral | No | Decreased GM areas | RNFL Temporal: Right eye= $66.66 \pm 14.07$ Left eye= $45.61 \pm 14.02$ |
| Frezzotti et al. 2014 | POAG | Bilateral | No | Decreased GM volume | IOP of poorer performing eye (mmHg) = $14.46 \pm 4.92$. IOP of better performing eye (mmHg) = $12.84 \pm 2.15$ |
| Frezzotti et al. 2016 | POAG | Unspecified | No | Decreased GM volume | RNFL of the poorer performing eye, mean±SD (μm) = $63.7 \pm 13.3$ |
| Giorgio et al. 2017 | POAG | Unspecified | No | Decreased GM volume | RNFL, mean±SD (μm) = $67.25 \pm 16.35$ IOP (on treatment), mean±SD (mm Hg) = $17.7 \pm 4.2$ |

GM, gray matter, HTG, high-tension glaucoma, IOP, intraocular pressure, LogMAR, logarithm of the minimum angle of resolution, NA, not available, NTG, normal tension glaucoma, POAG, primary open-angle glaucoma, RNFL, retinal nerve fiber layer, RNFLT, peripapillary retinal nerve fiber layer thickness, SD, standard deviation

**Table 2.**
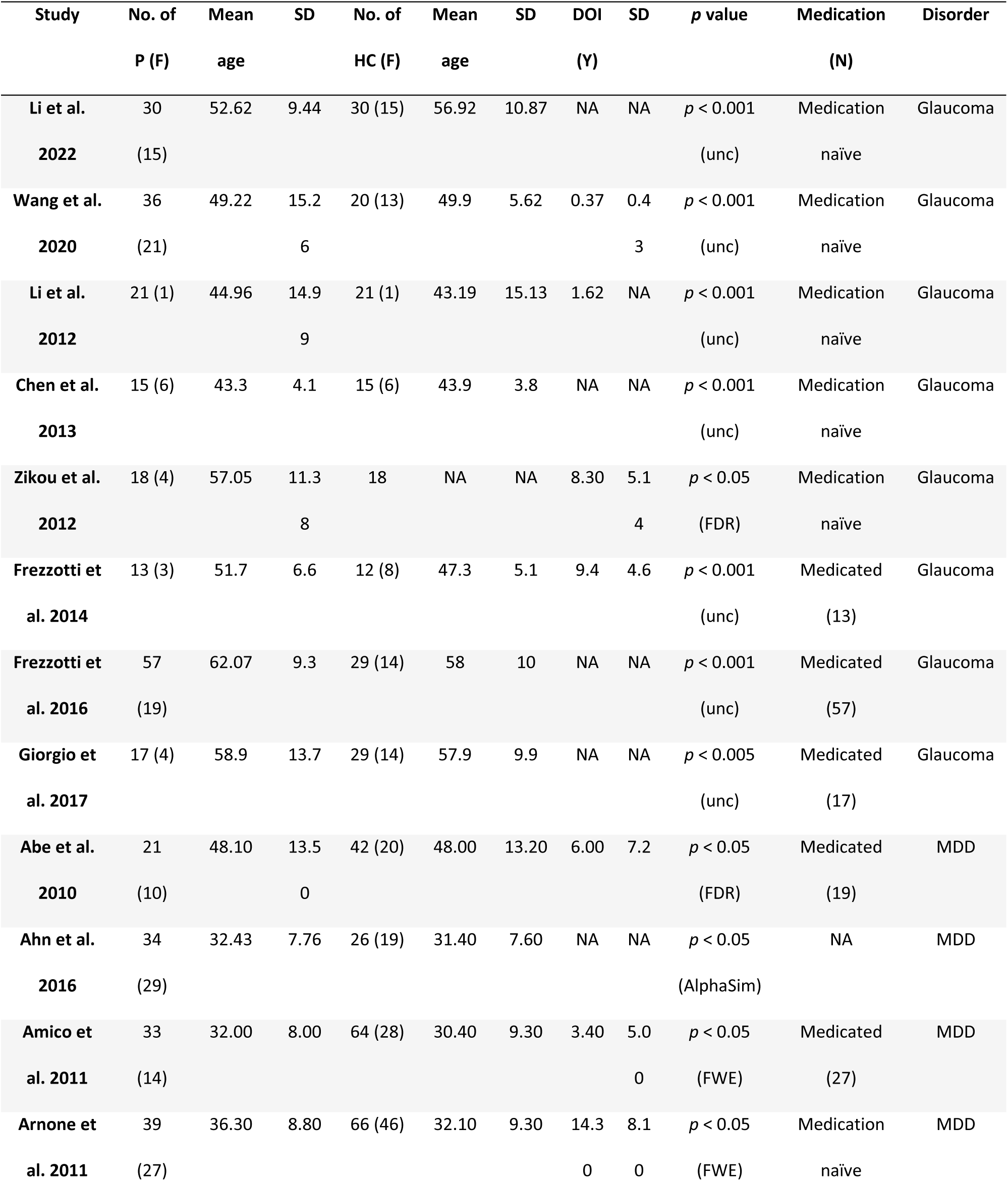
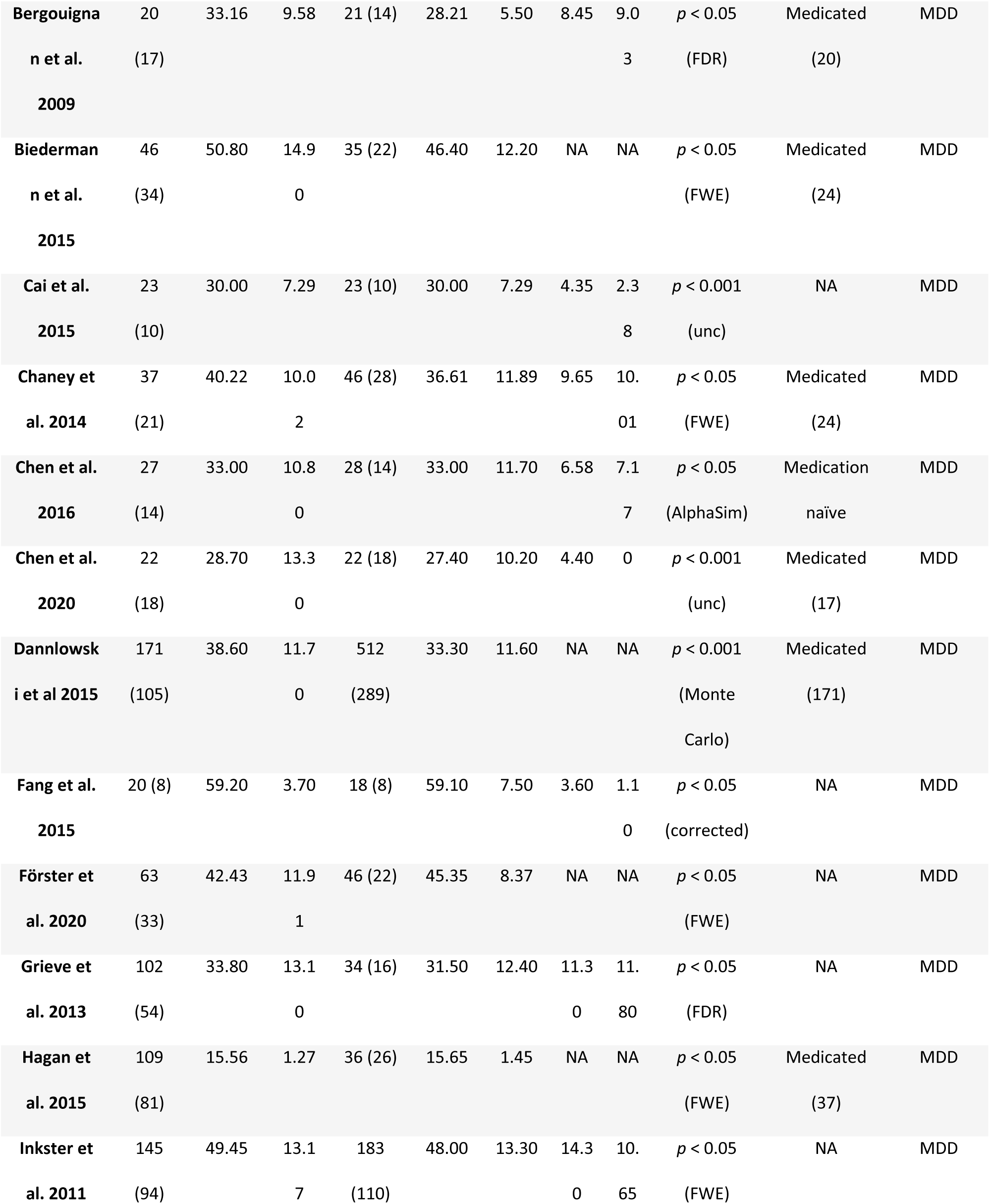
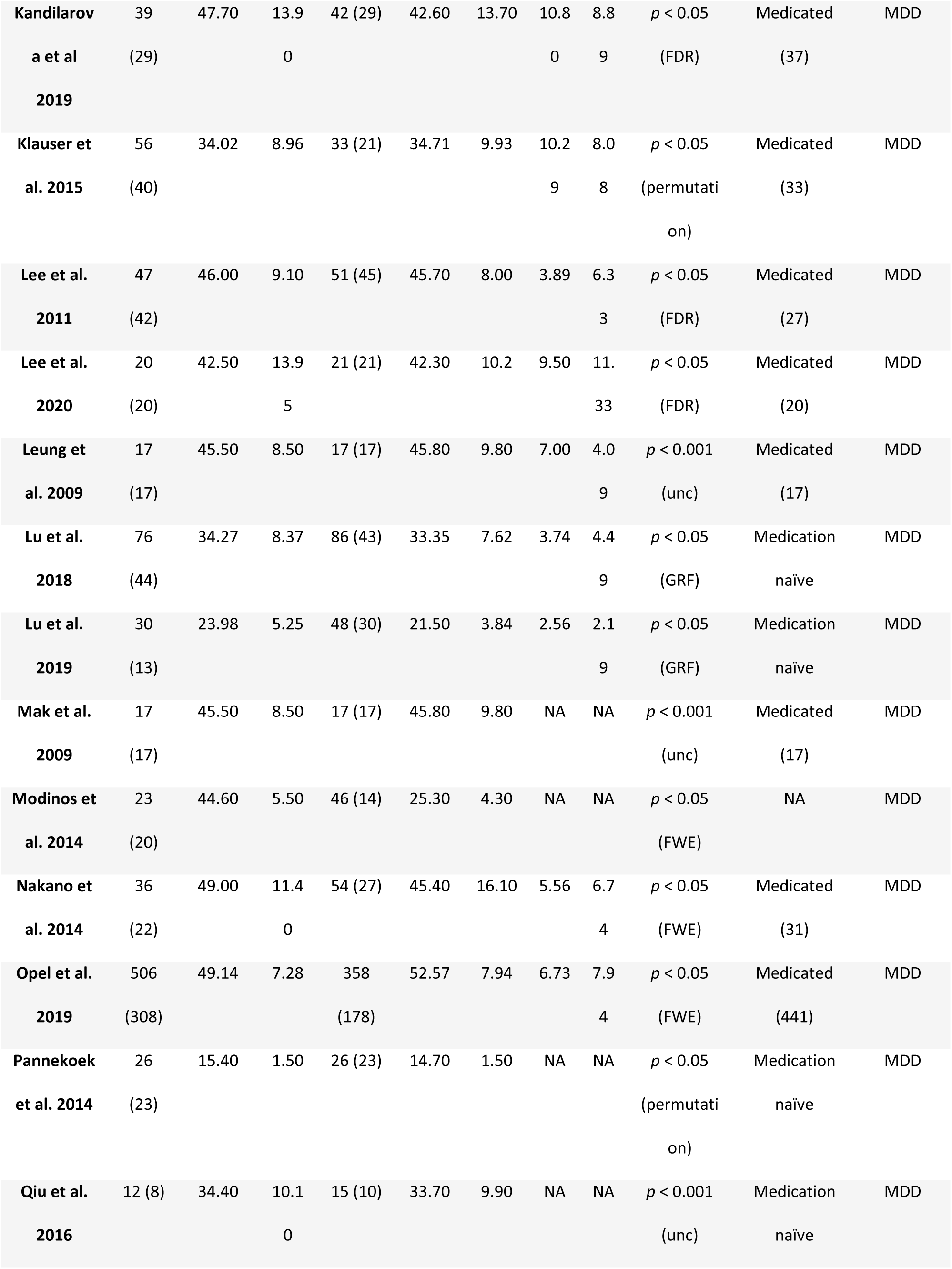
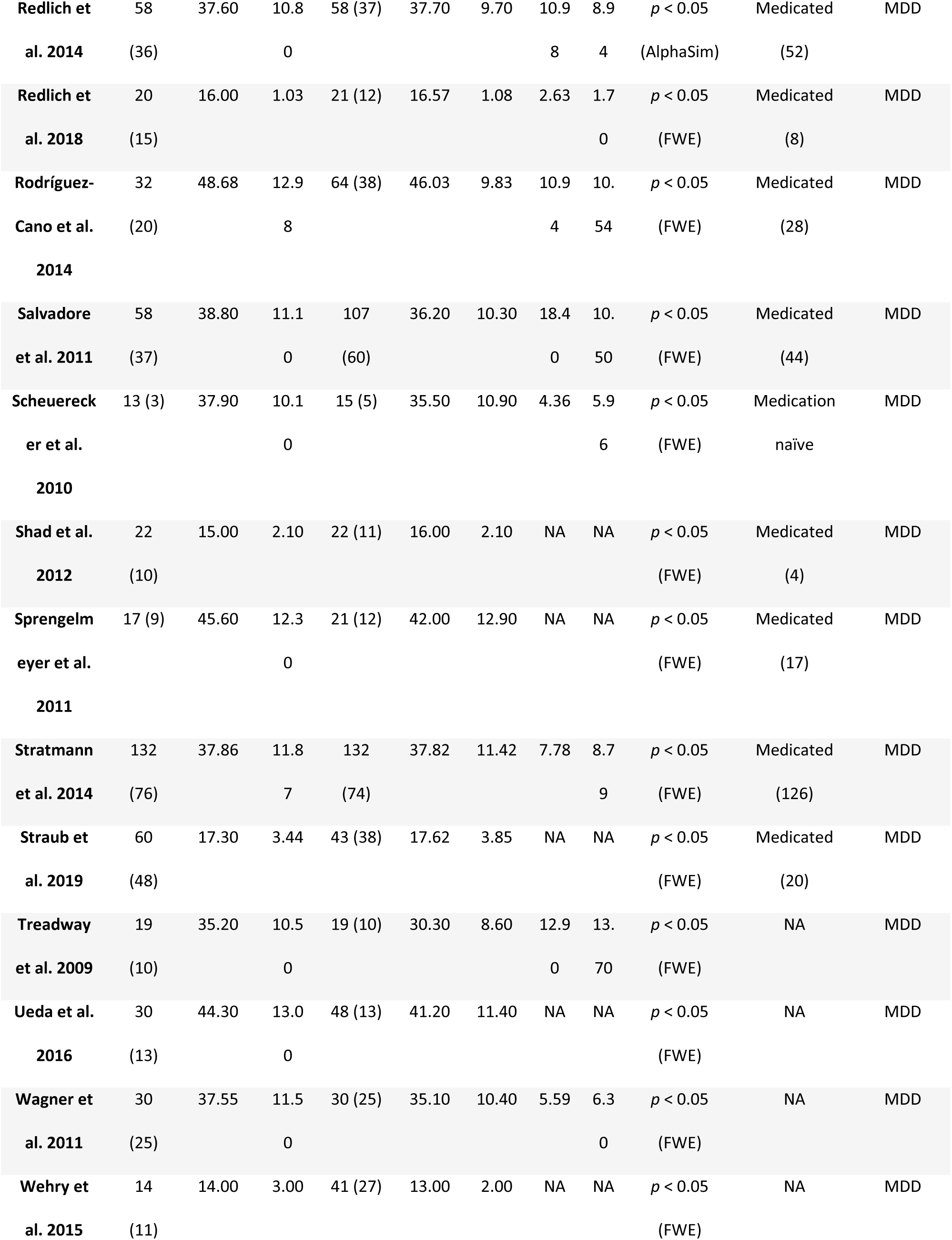
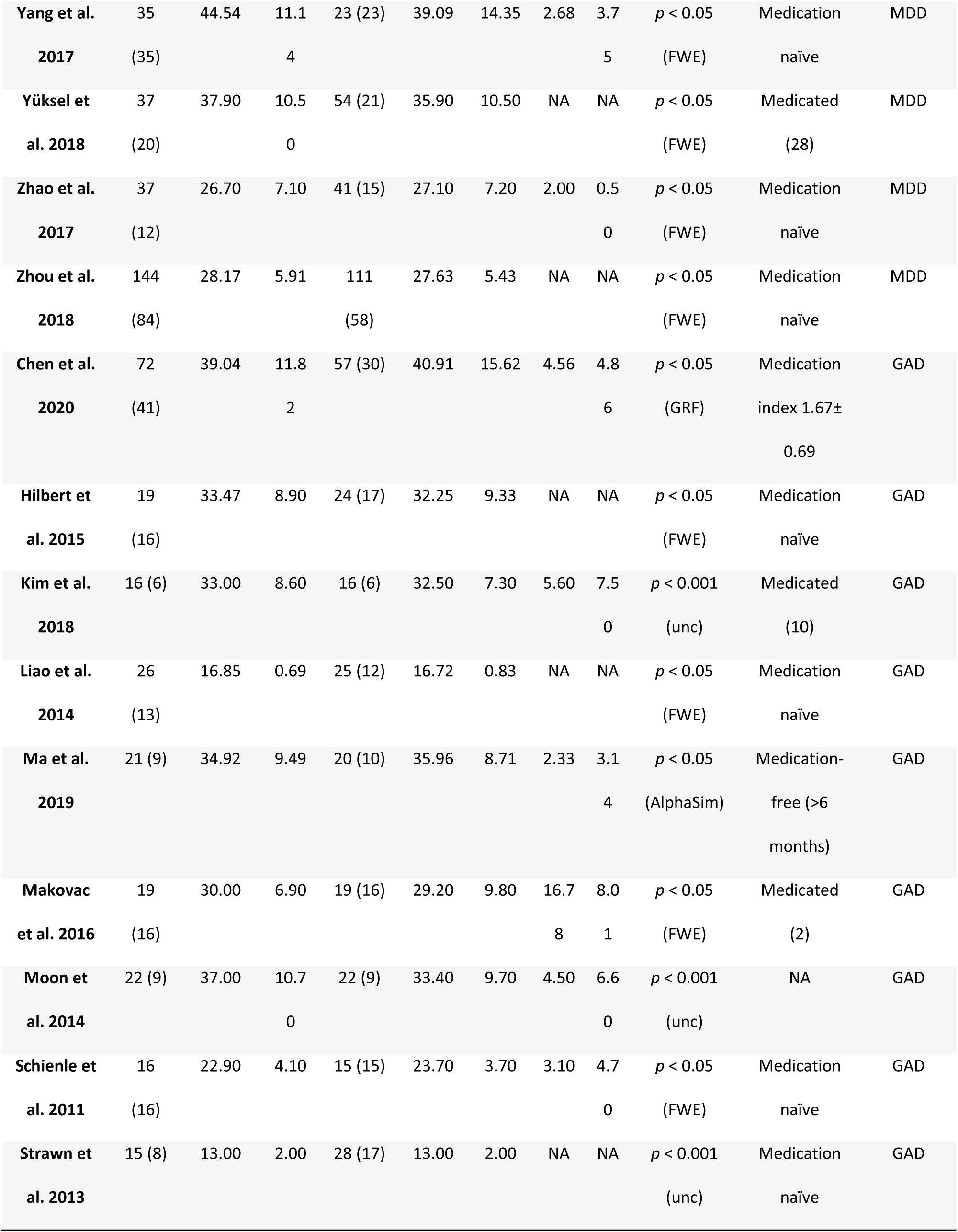
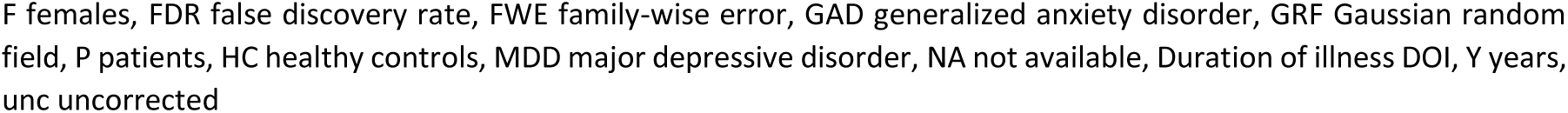
Demographic and clinical characteristics of the included datasets across the 3 disorders, Glaucoma, MDD and GAD.

### Regional gray matter alterations in glaucoma, GAD and MDD

The disorder-focused meta-analysis in glaucoma demonstrated robust and regional-specific GMV decreases in regions associated with visual and visuo-spatial processing, with significant clusters located in the left medial surface of the occipital cortex (lingual gyrus), the left thalamus and the left superior parietal lobe. Given that the thalamus is an anatomically and functionally heterogenous region, we further mapped the identified thalamic cluster using a high-resolution MRI-based atlas of the human thalamic nuclei (Saranathan et al., 2021)suggesting that the GMV decreases in glaucoma were located in the pulvinar. Glaucoma patients moreover displayed robust decreases in a number of other regions including the left insula/Rolandic operculum and putamen as well as the right precentral gyrus (Fig. 2A, details also reported in Table 3). Details of the separate meta-analytic results in GAD and MDD are reported in (X. Liu et al., 2022). Briefly, relative to the healthy controls GAD patients exhibited robust decreases in a network encompassing the left insula/Rolandic operculum, inferior frontal, thalamic and right inferior parietal regions and increased GMV in the right paracentral lobule (p < 0.0025, uncorrected) (Fig. 2B). MDD patients exhibited decreased GMV encompassing a network of the right insula/STG/Rolandic operculum, bilateral medial OFC and anterior cingulate cortex (ACC), as well as paracingulate gyri and right parahippocampal gyrus (p < 0.0025, uncorrected) (Fig. 2C).

**Figure 1:**
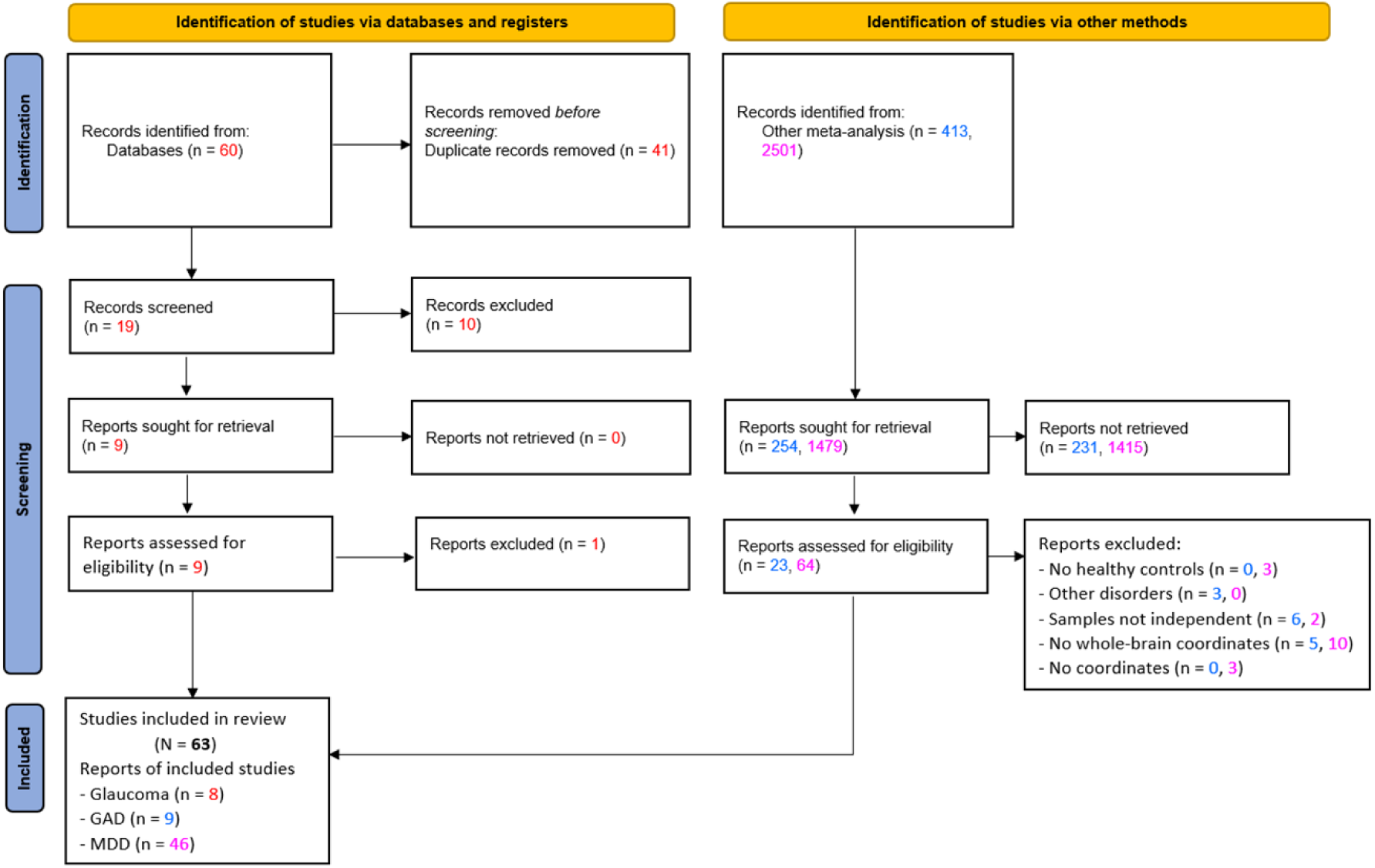
PRISMA flow diagram

**Figure 2.**
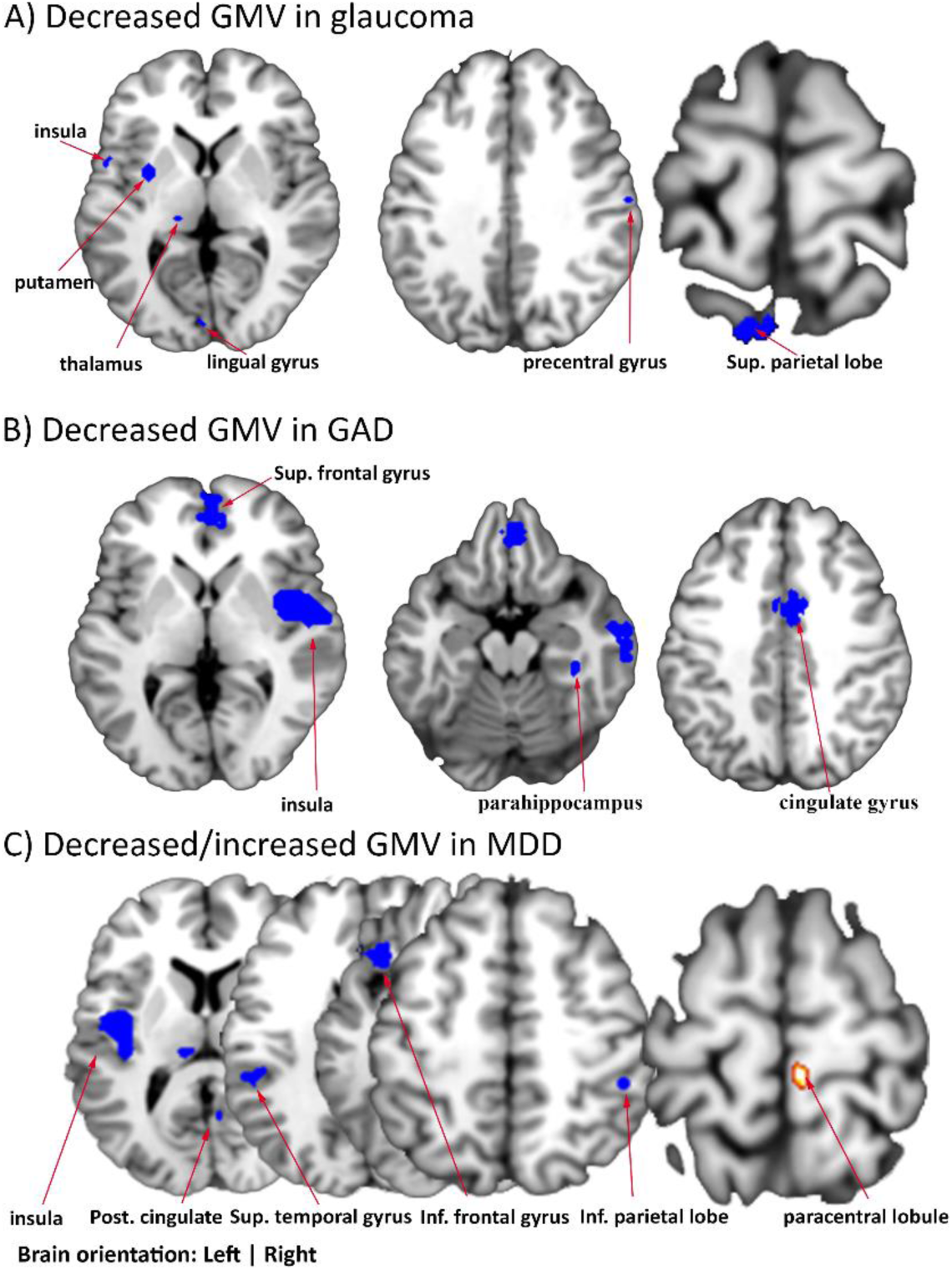
Gray matter decreases in glaucoma, GAD and MDD. A. Glaucoma: Decreases primarily observed in visual processing regions such as the left thalamus, lingual gyrus, and superior parietal lobe, as well as regions not primarily involved in visual processing such as the left insula/Rolandic operculum and right precentral gyrus. B. GAD: As per Liu et al. (2022), marked GMV decreases in left insula/Rolandic operculum, inferior frontal, thalamus; increase in right paracentral lobule (p < 0.0025, uncorrected). C. MDD: Decreased GMV in networks including right insula/STG/Rolandic operculum, bilateral medial OFC, ACC, and right parahippocampal gyrus (p < 0.0025, uncorrected).

**Table 3.**
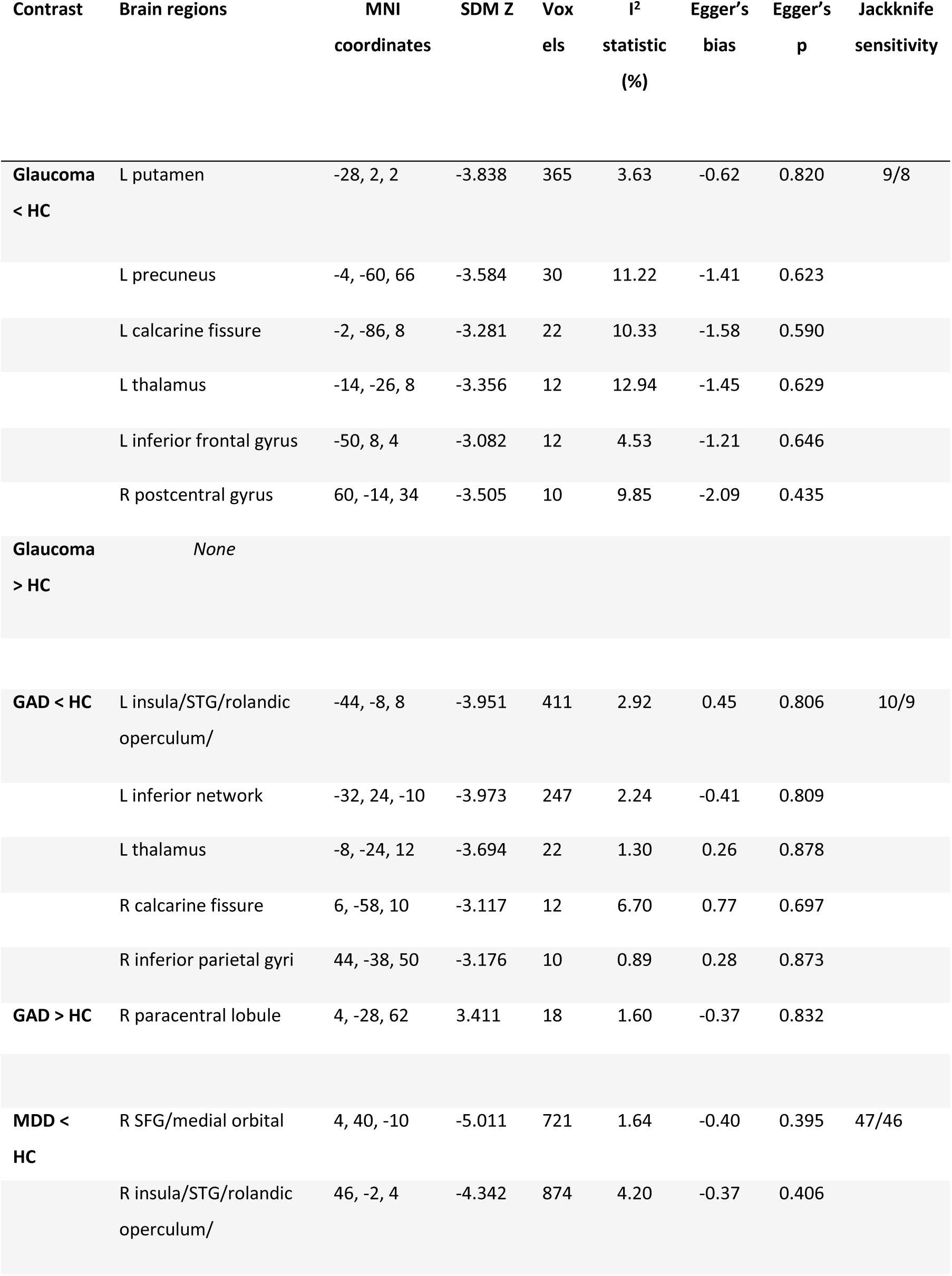
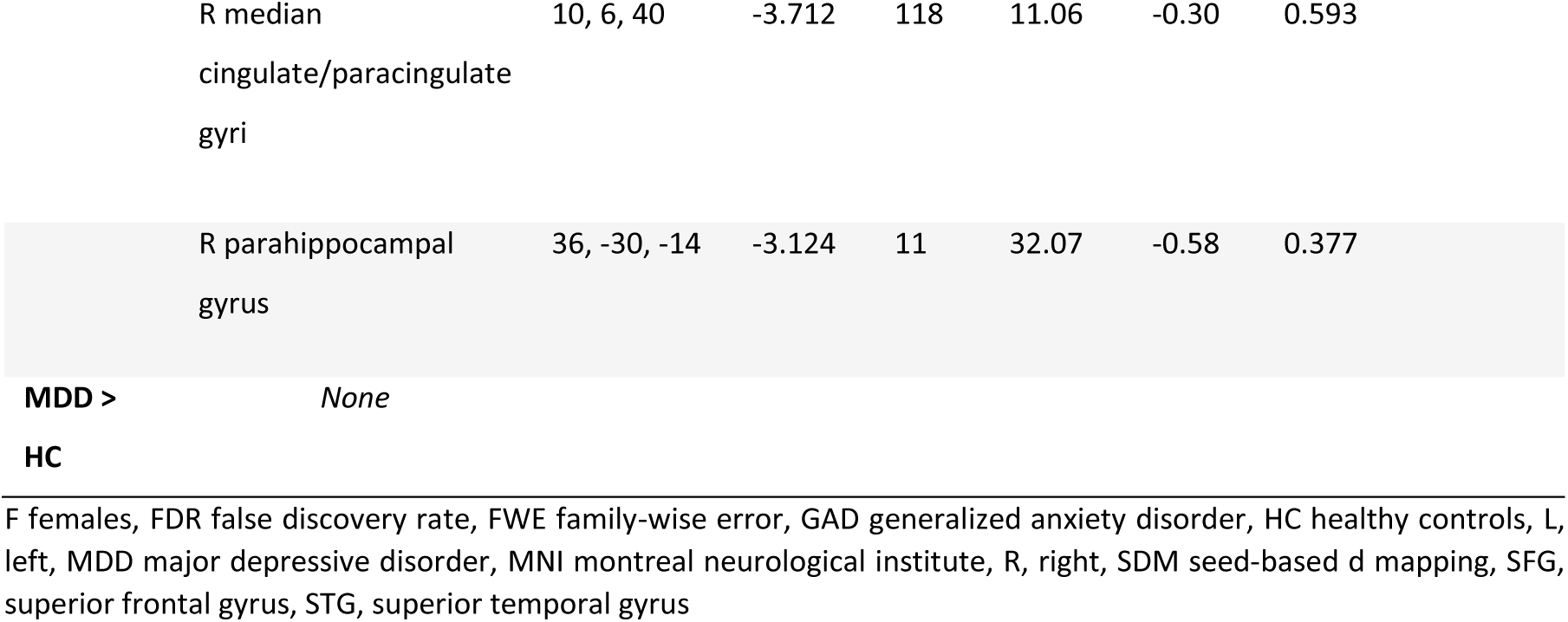
Whole-brain meta-analytic results of V Glaucoma, GAD and MDD.

### Sensitivity, heterogeneity and publication bias

The results of tests for heterogeneity, publication bias as well as sensitivity analyses are presented in table 3. The analysis of heterogeneity tests across disorders was generally low to moderate (ranging from 0.89% to 32.07%). Publication bias failed to reach statistical significance. The jackknife sensitivity analyses in the identified regions were highly replicable across all combinations.

### Meta regression for the glaucoma studies

Meta-regression with respect to age and or gender of the patients did not reveal significant effects of the meta-analytic results of glaucoma (Supplementary Figure 4 and 5).

### Functional characterization of the identified regions

We next aimed at determining whether the identified regions represent part of distinct large-scale functional systems and which behavioral domains are associated with these regions. We focused on an identified region primarily involved in visual processing (i.e., lingual gyrus) as well as two regions classically not and regions not primarily involved in visual processing (insula and putamen). Meta-analytic connectivity and co-activation analyses were employed (Fig. 3). Findings revealed that, the lingual region showed strong connectivity with visual networks, encompassing widespread regions of the occipital lobe and primary visual processing regions such as the calcarine, fusiform gyrus, and parts of the cuneus (Fig. 3A). For the left insula, the connectivity and co-activation patterns primarily encompassed the bilateral insular cortex and the dorsal anterior cingulate which together constitute the salience network. Additionally, the connectivity and co-activation patterns of the left putamen converged on a network encompassing bilateral dorsal and ventral regions strongly engaged in reward and motivational processes (Fig. 3C).

**Figure 3.**
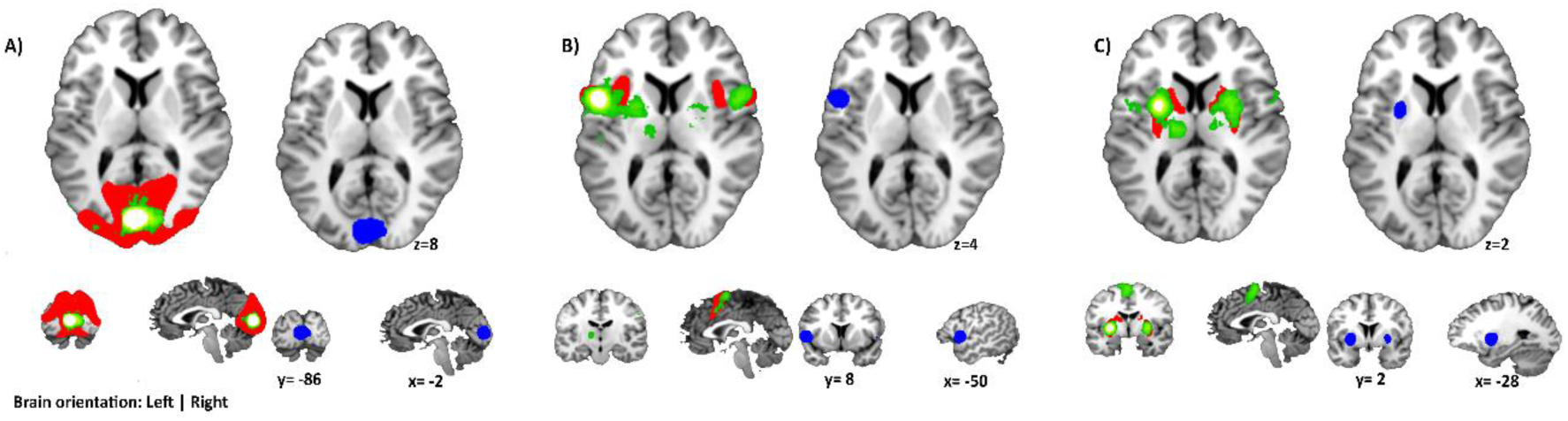
Meta-analytics co-activation and functional connectivity modeling of the regions found to be altered in glaucoma utilizing the neurosynth platform. (A) displays regions that exhibited a significant correlation with the lingual region, (B) displays regions exhibiting a significant correlation with the identified insular region, (C) shows regions that exhibited a significant correlation with the identified putamen region. The depicted regions were obtained from the Neurosynth database, with a significance threshold set at p-value = 0.01. In the figure, the red regions represent functional connectivity, green regions indicate meta-analytic coactivation, while the blue regions show the intersection between the functional connectivity and the meta-analytic co-activations

Behavioral characterization of the identified regions, including the lingual gyrus, putamen, and insula, was performed using the Neurosynth decoder. Analysis of the top ten behavioral terms revealed that the identified regions were associated with visual processing, pain processing and executive control as well as reward and motivation (Fig. 4). In particular, the lingual gyrus displayed a strong association with processes involved in visual processing and motion perception (r=0.3-0.35), while the insula and putamen exhibited a strong association with emotional/pain and reward related processes (r=0.25 and r=0.5), respectively.

**Figure 4.**
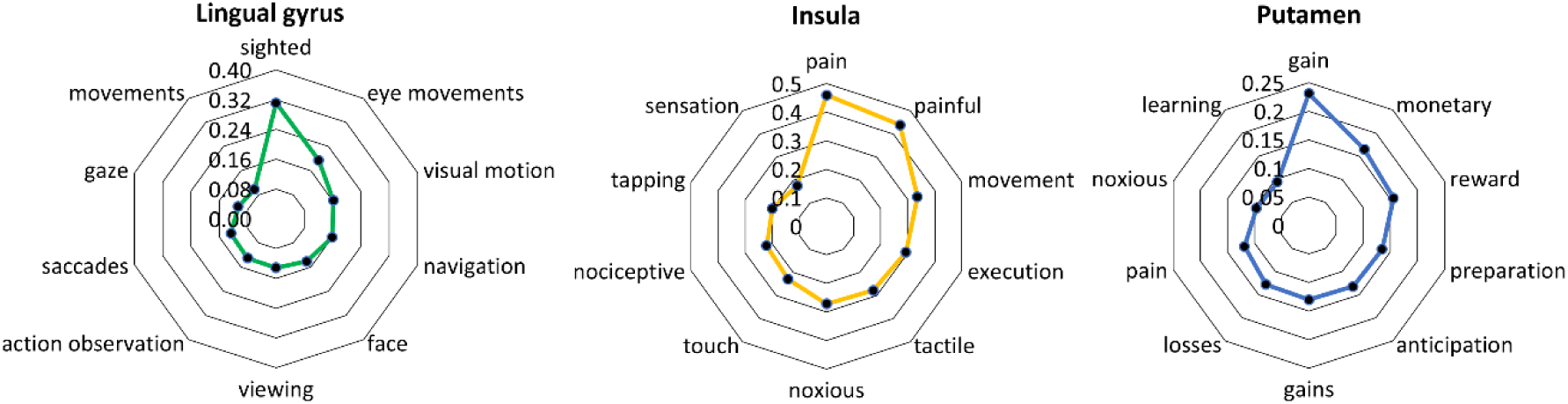
Functional characterization of the core regions obtained in the glaucoma meta-analysis. Each figure shows the top ten coefficients associated with the brain seed region.

### Overlap with anxiety and depressive disorders

For the conjunction analysis, we found that among the three disorders, only glaucoma and GAD exhibited an overlap in terms of GMV decrease in the left insula and the adjacent left precentral gyrus (Figs. 5 A and B), obtained with a multiple correction threshold at TFCE (p=0.05).

**Figure 5.**
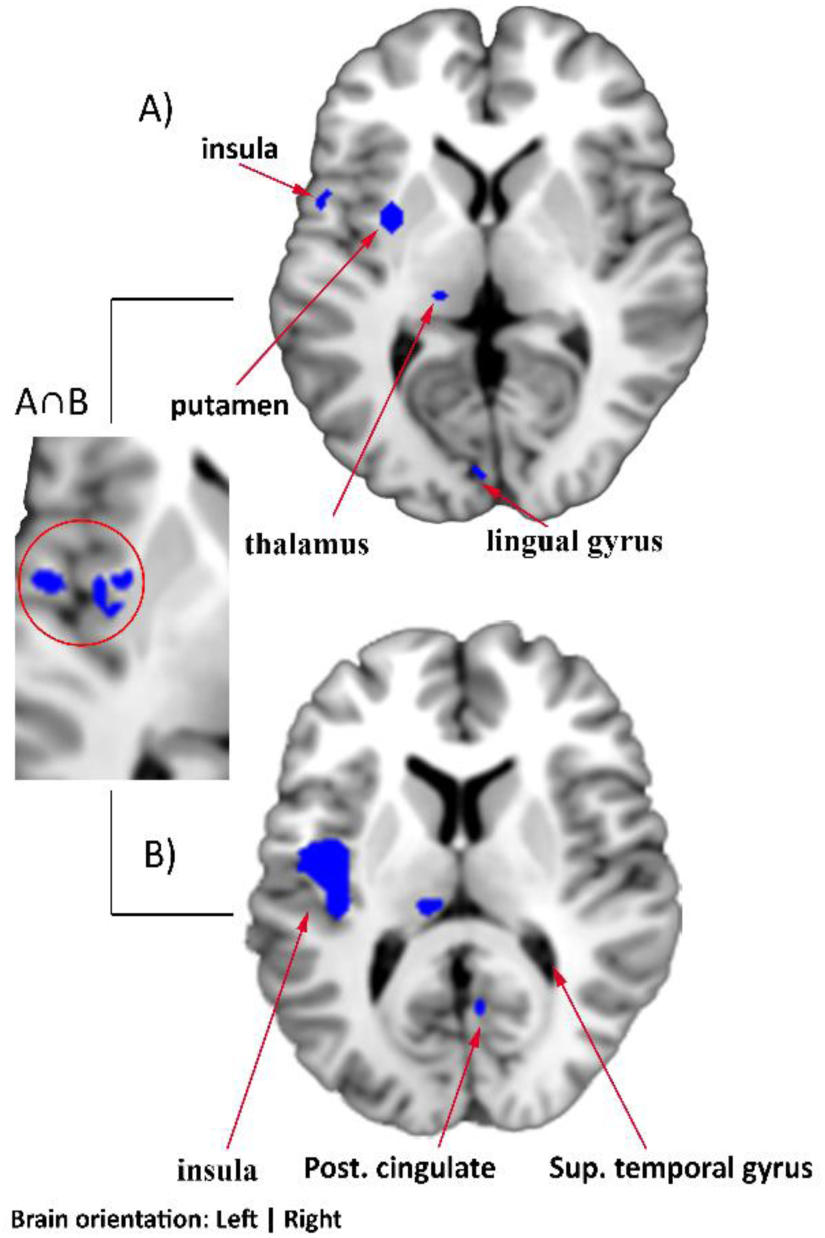
Conjunction analysis of GMV changes in glaucoma and GAD. Both disorders showed overlapping GMV decrease in the left insula and adjacent left precentral gyrus. Results were obtained using multiple correction thresholds with TFCE (p=0.05).

## Discussion

With an estimated 79.6 million individuals affected, glaucoma has become the leading cause of irreversible blindness (Quigley & Broman, 2006; Tham et al., 2014). Despite a growing number of qualitative reviews and perspectives suggesting that glaucoma is accompanied by brain structural alterations within and beyond the visual system (e.g. (Chan et al., 2021; Nuzzi et al., 2018; H. J. Zhang et al., 2019)), a systematic determination of robust neurostructural brain alterations in glaucoma is lacking. Here we capitalized on the growing number of studies employing MRI-based structural neuroimaging in glaucoma patients. A quantitative evaluation of the published findings revealed robust gray matter reductions in a left lateralized network encompassing regions involved in visual processing such as the medial surface of the occipital lobe (lingual gyrus), the pulvinar and the left superior parietal lobule. Moreover, volumetric reductions were observed in regions that are commonly not considered as primary visual processing regions such as the left insula and putamen. The hypothesized impact on brain systems that reach beyond the visual system was further confirmed by results from network level and behavioral characterization analyses. The corresponding results indicated that the identified lingual gyrus region was part of a functional network encompassing the primary and secondary visual cortex and was associated with visual and oculomotor behavioral domains. In contrast, the insular region exhibited strong functional interactions with the salience network encompassing the bilateral anterior insula and dorsal anterior cingulate cortex and was behaviorally characterized by terms referring to pain and negative emotional experiences. The identified putamen region exhibited functional interactions with the entire striatal system as well as a behavioral characterization revolving around reward and motivational processes. Based on the high prevalence rates of depression and generalized anxiety in glaucoma patients, we further explored overlapping alterations with meta-analytic gray matter maps of these disorders and observed that generalized anxiety and glaucoma share gray matter reductions in the left insular cortex. Together, our results confirm gray matter reductions in glaucoma that affect visual processing systems as well as insular and striatal systems involved in affective and reward-related processes.

The main meta-analysis revealed robustly decreased gray matter volume in patients with glaucoma as compared to healthy controls in a set of regions involved in visual processing. While no robust changes were found in the primary visual cortex, we found decreased gray matter in the left thalamus, lingual gyrus and superior parietal lobule. The thalamus is a functionally and anatomically heterogenous region that plays crucial roles in visual processing. The lateral geniculate nucleus of the thalamus relays visual information from the retina via the optic nerve and projects this information to the primary visual cortex. Further mapping of the identified region using a high-resolution atlas revealed that the identified region was located in the pulvinar. The pulvinar is the largest nucleus in the thalamus and exhibits extensive and reciprocal interactions encompassing subregion-specific connections with primary and extrastriate visual processing areas as well as cortical regions, in particular parietal and frontal regions (Arcaro et al., 2015). While this central position supports the involvement of the pulvinar in visual attention (e.g.(Fiebelkorn & Kastner, 2020)) accumulating evidence additionally suggests a critical role of this region in basic visual processing such that deactivation of the pulvinar in non-human primate models causes strongly decreased sensory responses in the visual cortex (see (Fiebelkorn & Kastner, 2019; H. Zhou et al., 2016). Pathological gray matter reductions additionally affected further systems engaged in different later visual processing stages, including the lingual gyrus located on the medial surface of the occipital cortex or the superior parietal lobule. Both regions exhibit strong reciprocal connections with the primary visual cortex (Palejwala et al., 2021) and contribute to higher visual processing operations including object recognition or the integration of information from primary visual processing areas with sensory and cognitive processes, respectively. The role of deficient lingual gyrus integrity in glaucoma is further corroborated by other imaging modalities, such that surface-based morphometry studies reported decreased lingual gyrus thickness (Nuzzi et al., 2018) and a recent functional MRI study reported that decreased network communication of this region associated with the extend of visual field loss (Demaria et al., 2022) in glaucoma patients.

The primary meta-analysis moreover indicated robust gray matter reductions in the left putamen and left insular cortex, and thus regions with no central or direct role in visual processing. In line with the results from the meta-analytic network and behavioral characterization, the putamen exhibits strong connections with other parts of the striatum and frontal regions and plays a crucial role in reward, motor and cognitive control domains (Haber, 2016; Kreitzer & Malenka, 2008; Zhao et al., 2019, 2021; Zhuang et al., 2021). Pathological alterations in this region have been reported across a range of neurodegenerative and mental disorders, including decreased gray matter volume in disorders associated with motor impairments such as Parkinson’s and Huntington’s Disorder (Minkova et al., 2017) or disorders associated with dysregulated reward and control processes i.e. addiction and obsessive compulsive disorder (Klugah-Brown et al., 2021; X. Zhou et al., 2020). The insula was identified as further system with robust gray matter decreases in glaucoma and the identified insular region connected strongly with the salience network centered around the bilateral insula and the dorsal ACC. In line with previous characterizations of the insula the region was associated with pain, interoceptive and salience processes (Ferraro, Klugah-Brown, Tench, Bazinet, et al., 2022; Ferraro, Klugah-Brown, Tench, Yao, et al., 2022; Uddin, 2015; F. Zhou et al., 2020). Decreased gray matter volume of the insula has been reported across a number of neurological and mental disorders, including migraine or mood disorders, and may represent a transdiagnostic markers for increased emotional distress (Goodkind et al., 2015; Taylor et al., 2023; Wise et al., 2017; X. Zhang et al., 2023). In our present data, the insula deficit in glaucoma patients is the only region overlapping with alterations as determined in our previous meta-analysis (X. Liu et al., 2022). Together these findings may indicate that reduced insular morphological integrity could represent a marker for emotional dysregulations or long-term stress exposure.

Our findings indicate that glaucoma is accompanied by robust alterations in a number of brain systems that play important roles in visual processing, such as the lingual gyrus or pulvinar, which may represent the neurostructural basis for the visual deficits in the patients. Moreover, alterations in the putamen – a region characterized by widespread interactions with the entire striatal system and reward-related behavioral domains – were observed which may neurally gate motivational or reward deficits in the patients. Finally, alterations in the insular cortex – in particular a regions communicating with the salience network and behaviorally associated with negative affective states – were observed and overlapped with GAD, which may reflect alterations related to enduring distress.

Several limitations need to be considered when interpreting the findings of this meta-analysis. First, the number of studies from the glaucoma and GAD cohorts included in this analysis is limited because there were few independent studies that met our inclusion criteria. Second, the inherent cross-sectional design focusing on structural MRI differences at a single time point limits our understanding of the chronological progression of neurostructural changes in glaucoma patients. A longitudinal study design may provide a more comprehensive understanding in future research. Although we found similarities in affected brain regions between glaucoma and emotional disorders, it is difficult to establish a direct or even cause-and-effect relationship. It remains to be determined whether the common neurostructural changes are a result of glaucoma directly leading to these emotional disturbances or whether they are influenced by other external or confounding.

## Data Availability

All data produced in the present study are available upon reasonable request to the authors

## Acknowledgements and funding

The study was supported by the National Natural Science Foundation of China (Grant No. 82271583, Grant No. 32250610208 to BB), and the National Key Research and Development Program of China, Grant No. 2018YFA0701400. Disclaimer: Any opinions, findings, conclusions, or recommendations expressed in this publication do not reflect the views of the Government of the Hong Kong Special Administrative Region or the Innovation and Technology Commission.

