## Supplementary Information for "The neurostructural consequences of glaucoma and their overlap with disorders exhibiting emotional dysregulations: a voxel-based meta-analysis and tripartite system model"

**Meta-regression**

We performed exploratory meta-regression analyses for age and gender effects across all clusters identified in the meta-analysis. In line with(Liu et al., 2022) potential confounding effects of demographic as well as clinical variables were examined in a meta-regression. Although we report the results for n ≥ 8 (Glaucoma studies),(Radua & Mataix-Cols, 2009) recommends variables reported in n ≥ 9 studies for a standard analysis.

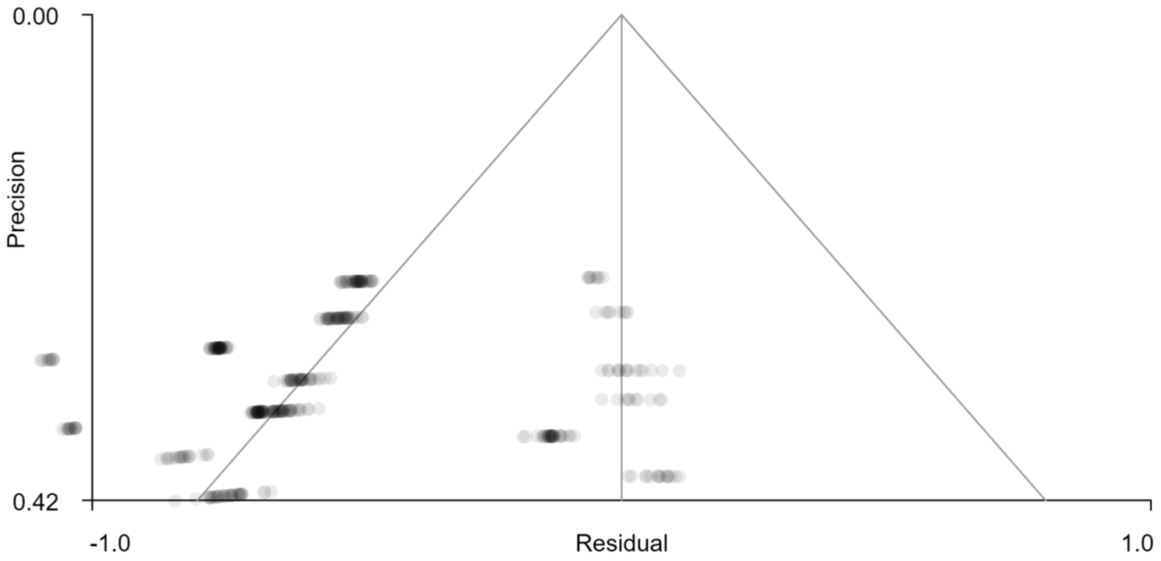


Supplementary Figure 1 Funnel plot for Glaucoma meta-analysis (L Putamen, MNI = -28, 2, 2)

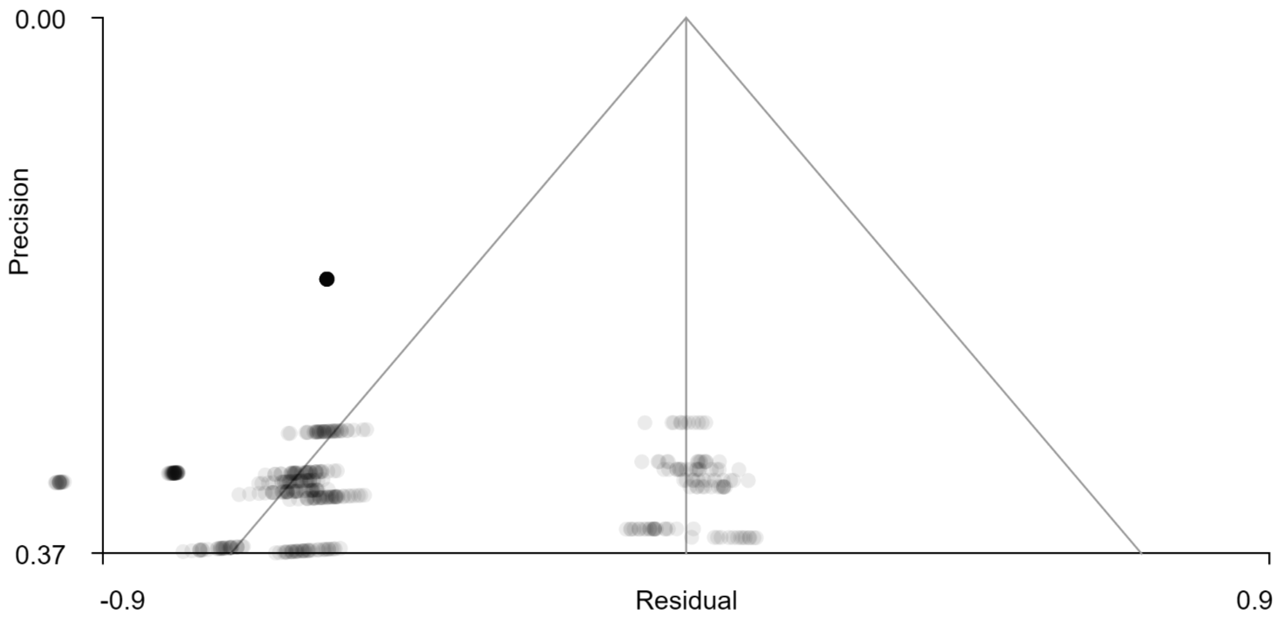


Supplementary Figure 2 Funnel plot for GAD meta-analysis (L rolandic operculum/Insula, MNI = -44, -8, 8)

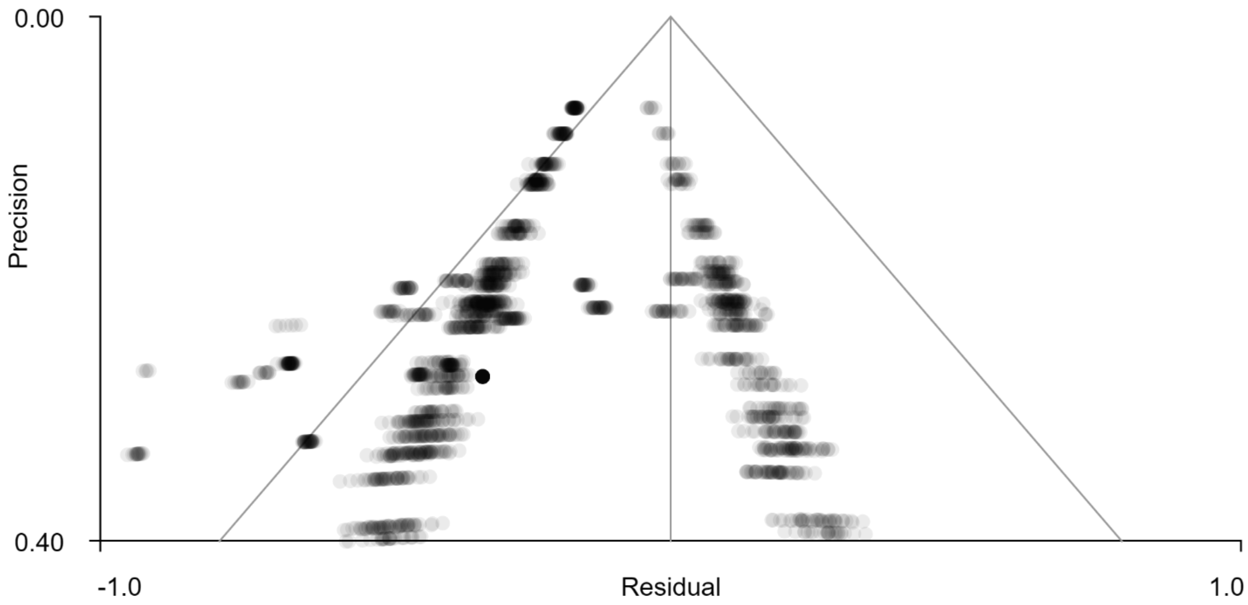


Supplementary Figure 3 Funnel plot for MDD meta-analysis (Superior frontal gyrus, MNI = 4, 40, -10)

### Meta regression analysis

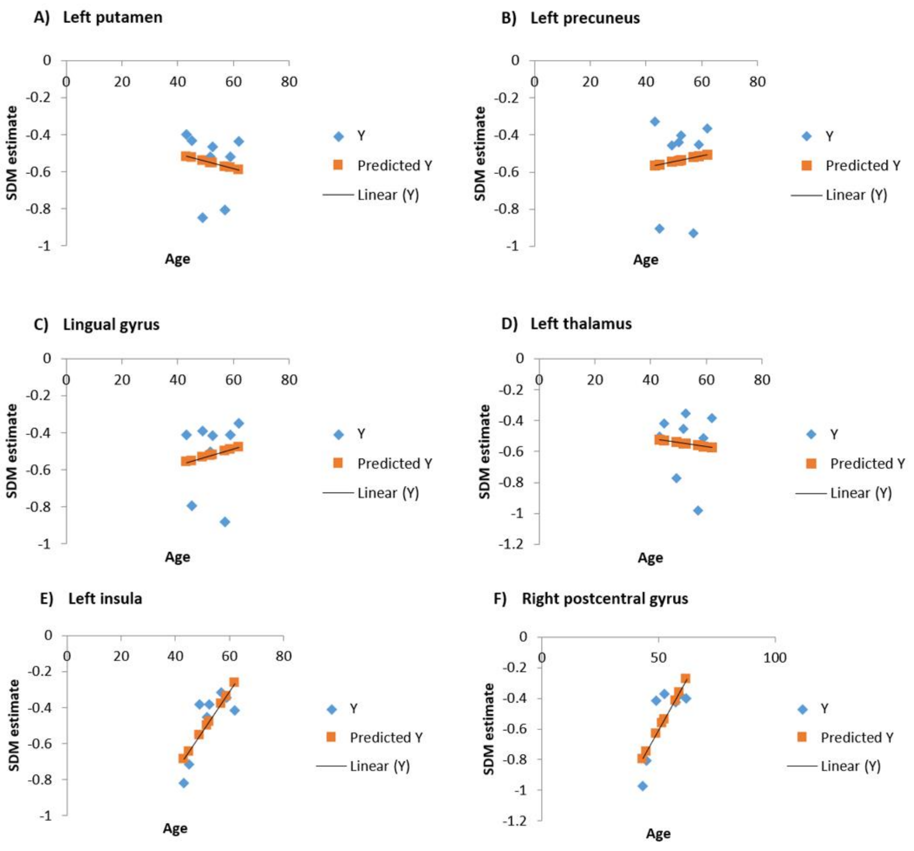


Supplementary Figure 4 Meta regression analysis between age and the meta-analytic results of glaucoma

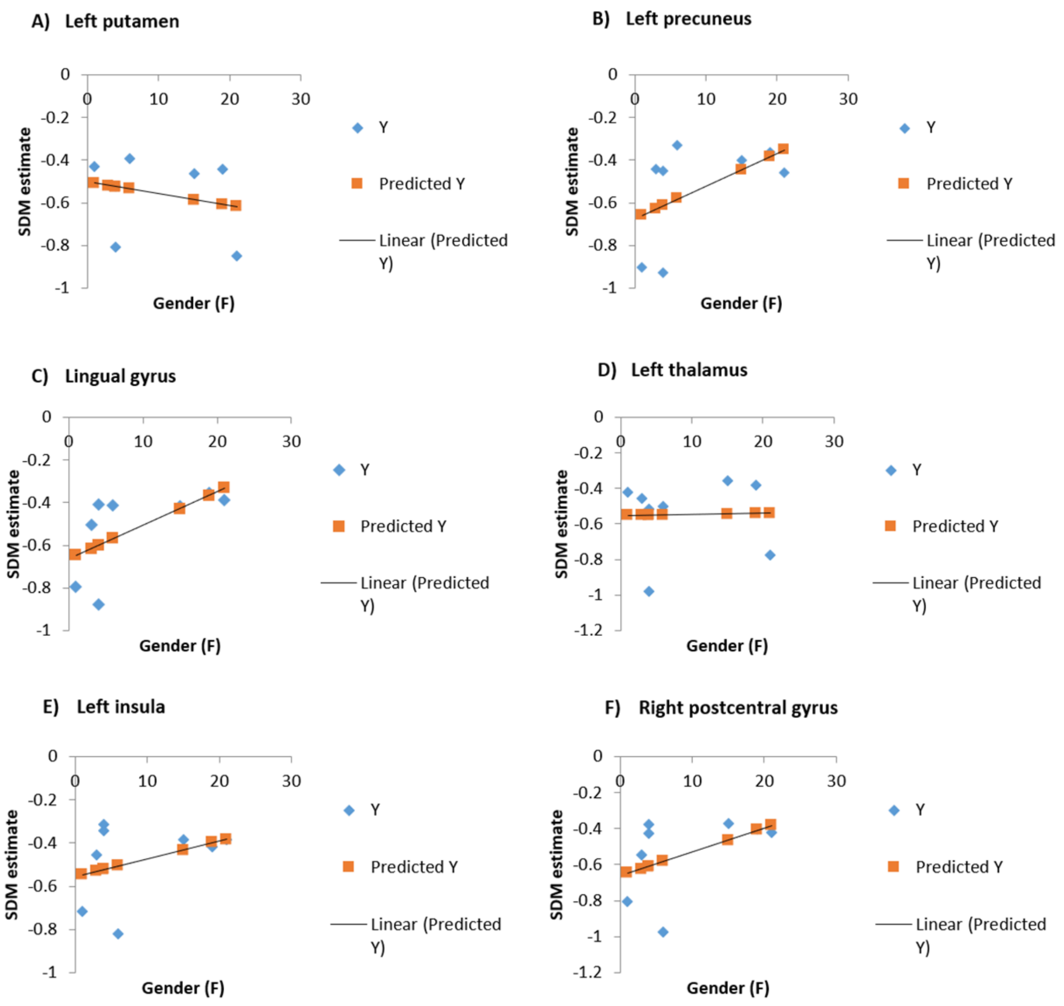


Supplementary Fig 5 Meta-regression between gender and the meta-analytic results of glaucoma. F, Females
